# Coronary Artery anomalies in PatienTs Undergoing transcatheter aortic valve REplacement. The CAPTURE registry

**DOI:** 10.1101/2023.08.15.23294143

**Authors:** Omar A. Oliva, Lorenzo Scalia, Antonio Sisinni, Enrico Poletti, Jessica Zannoni, Mattia Mazzucca, Enrico Criscione, Antonio Popolo Rubbio, Riccardo Gorla, Mauro Agnifili, Francesco Bedogni, Luca Testa

**Affiliations:** Department of Clinical and Interventional Cardiology, IRCCS Policlinico San Donato, 20097 San Donato Milanese (MI), Italy

**Author notes:** Address for correspondence*: Luca Testa, MD, PhD, Department of Clinical and Interventional Cardiology, IRCCS Policlinico San Donato, Piazza Edmondo Malan 2, 20097 San Donato Milanese (Milano), Italy.

**Keywords:** Coronary artery anomalies, TAVI, bioprosthesis, aortic valve replacement, aortic stenosis

## Abstract

**Background:** The incidence and possible implications of coronary artery anomalies (CAA) in patients undergoing transcatheter aortic valve replacement (TAVR) are uncertain.

**Aims:** To evaluate the impact of CAA on TAVR outcomes, and to suggest possible strategies to prevent CAA related complications.

**Methods:** Among 2,164 consecutive patients who underwent TAVR in our center, 15 (0.69%) were identified to have a CAA, based on pre-operative Computed Tomography (CT) scans.

**Results:** CT-angiography revealed the following features of CAA: the majority of anomalous CAA concerned the right coronary artery (80%), followed by the left main (13.3%) and the left circumflex (6.7%). An intramural course was observed only in 26.7% patients, whereas an acute take-off was detected in more than half of the study cohort (53.3%). High-risk sudden cardiac death features were observed in 6 (40%) patients.

Technical success was 86.7%, device success was 80%. One patient experienced a cardiac arrest 15 minutes after procedure, resulting from occlusion of the anomalous right coronary artery with the ostium located at the right-to-non coronary commissure. There were no cases of ≥ moderate paravalvular leak or stroke. One non-cardiac related death was recorded 4 months after the procedure.

**Conclusions:** The interaction between transcatheter bioprosthesis and different CAA types could lead to ominous sequelae, if not promptly recognized and treated accordingly.

## Introduction

Coronary artery anomalies (CAA) are rare findings, often incidentally encountered. The most common phenotype is an anomalous origin from the aorta,^1^ occurring in approximately 0.8% of patients undergoing coronary artery angiography. However, if we exclude cases where the left anterior descending (LAD) and left circumflex artery (LCx) have separated origins from the left sinus of Valsalva (LSV), the incidence drops to 0.2%.^2^ The assessment of the risk of sudden cardiac death and subsequent treatment is still a matter of debate due to limited data availability.^3^ Nevertheless, there are few anomalies that could be injured during surgical aortic valve replacement (SAVR), mainly LCx from right sinus of Valsalva (RSV) with a retro-aortic course.^4,5^

In this variant, the peri-procedural is determined by the prosthesis compression or suture ligation.^6^ Surgeons have implemented different precautions to ensure the safety of surgical treatment, including the following: i) undersizing of the aortic valve bioprosthesis;^5,7,8^ ii) dissection of the LCx from the aortic annulus;^8–10^ iii) tilting of the bioprosthesis;^11^ sutureless bioprosthesis, thanks to ligation risk absence and to inferior impact on the aortic anulus;^12^ iv) supranular or stentless bioprosthesis implantation^5^ and/or v) revascularization only in case of overt ischemia, through coronary artery bypass grafting^4,13^ or drug eluting stent implantation.^6,13^

Other anomalies can be treated with transection and re-implantation (e.g. RCA from LSV with retro-aortic course)^14^ or with remodelling of the adjacent structure (e.g. pulmonary trunk and right ventricle) to avoid compression (e.g. RCA from LSV with an inter-arterial course). ^15^

Transcatheter aortic valve replacement (TAVR) has been established as an effective treatment for patients with severe aortic stenosis ^16^. Since the number of TAVR cases is meant to grow with the expansion of the indication to younger and lower risk patients, the likelihood of performing TAVR in the presence of a CAA should consistently increase.

The aim of the study is to evaluate the incidence and clinical implications of CAAs in patients undergoing TAVR, and to propose possible management algorithms.

## Methods

### Study design and data collection

This observational retrospective single-center study analyzed all consecutive patients with evidence of CAA who underwent TAVR procedure from July 2015 to March 2023 at our Center.

Inclusion criteria were: 1-detection of CAA at pre-TAVR computed tomography (CT) scan; 2-patient with severe aortic stenosis of a native valve or degenerated bioprosthesis with an indication for transcatheter treatment. Exclusion criteria was previous cardiac surgery for CAA re-implantation.

Data regarding baseline clinical and echocardiographic characteristics, CT scan, procedural features, as well as follow-up at 30 day and 1 year, were collected into our electronic prospective institutional database.

Patients were deemed suitable for TAVR after Heart Team evaluation, and they all provided written informed consent for the procedure.

The study complied with the Declaration of Helsinki. The local ethics committees approved the use of retrospective anonymized data for this study.

### CT scan and transthoracic echocardiography

CT-angiography scans were performed on 64- or 128-row electrocardiography gated multidetector scanner (Somatom Definition; Siemens healthcare, Forchheim, Germany). The 3-Mensio valves software (version 8.2, Pie Medical Imaging, Maastricht, The Netherlands) enabled multiplanar reconstruction analysis of the aortic root and of the coronary arteries, evaluating both the diastolic and systolic phases.^17^ The following features of coronary arteries were evaluated: i) take-off level, below or above aortic valve commissure; ii) take-off angle, acute (<45°) or non-acute (≥ 45°); iii) course, pre-pulmonic, inter-arterial, sub-pulmonic, retro-aortic or retro-cardiac; iv) ostia type, separate, shared or branch vessel; v) proximal tract morphology, normal, oval or slit-like and vi) evidence of intramural location, as suggested by Cheezum et al.^18^, allowing to plan a strategy to avoid negative interaction with TAVR devices. Measurements were made by a dedicated laboratory of radiology technicians.

Oversizing was determined as: calculated perimeter oversizing (%) = [(prosthesis perimeter/CT valve perimeter -1) *100].

Transthoracic echocardiography (TTE) was performed pre- and post-procedural, the latter repeated at discharge. Post-procedural paravalvular leak (PVL) was arranged evaluated by experienced echocardiographers, according to Valve Academic Research Consortium-3 (VARC-3) criteria.^19^

### TAVR procedure

Transfemoral TAVR was performed under local anesthesia. Conscious sedation was added according to patient’s tolerance. The type of bioprosthesis was selected by the first operator based on the CT scan images.

### Study end-points

Technical success was defined in accordance with VARC-3 criteria as: i) freedom from mortality; ii) successful access, delivery of the device, and retrieval of the delivery system; iii) correct positioning of a single prosthetic heart valve into the proper anatomical location and iv) freedom from surgery or intervention related to the device (permanent pacemaker implantation excluded), or to a major vascular or access-related, or cardiac structural complication.

Device success at 30 days was referred to the attainment of technical success and the fulfillment of the following situations: i) freedom from mortality; ii) freedom from surgery or intervention related to device (permanent pacemaker implantation excluded), major vascular, access-related or cardiac structural complication; and iii) intended performance of the valve, as mean gradient <20 mmHg, peak velocity <3 m/s, Doppler velocity index ≥0.25, and less than moderate aortic regurgitation.^19^

### >Statistical analysis

Categorical and dichotomous variables are displayed as frequencies and percentages, meanwhile, continuous variables are reported as mean and standard deviation, or median and interquartile range, as appropriate.

Analyses were performed using SPSS statistical analysis software version 28.0 (IBM Corporation, Armonk, NY, USA).

## Results

### Baseline characteristics

Among the last 2,164 patients treated by means of TAVR, only 15 (0.69%) were identified to have a CAA on pre-operative CT scan.

The mean age was 83.7 ± 7.9 years, and the majority were female (80.0%). Mean STS score was 4.0 ± 2.8. One patient (ID 10) had prior SAVR with a Trifecta 23 bioprosthesis, meanwhile 2 patients (20%) suffered of coronary artery disease. Of those, one (ID 2) had previous coronary artery bypass surgery with evidence of occlusion of the saphenous graft for the anomalous RCA, whereas the other one (ID 7) suffered from a chronic total occlusion of the anomalous RCA due to intrastent restenosis in the middle and proximal tract.

CT-angiography identified CAA features: most anomalies affected RCA (80%), with LM (13.3%) and LCx (6.7%) to follow. Intramural course was observed only in 4 patients (26.7%), meanwhile, acute take-off was detected in more than half (53.3%) of the study cohort. Ostia with high-risk sudden cardiac death features (oval or slit like) were observed in 6 patients (40%), as shown in ***Table 1 and Suppl Table 1***.

**Table 1.**
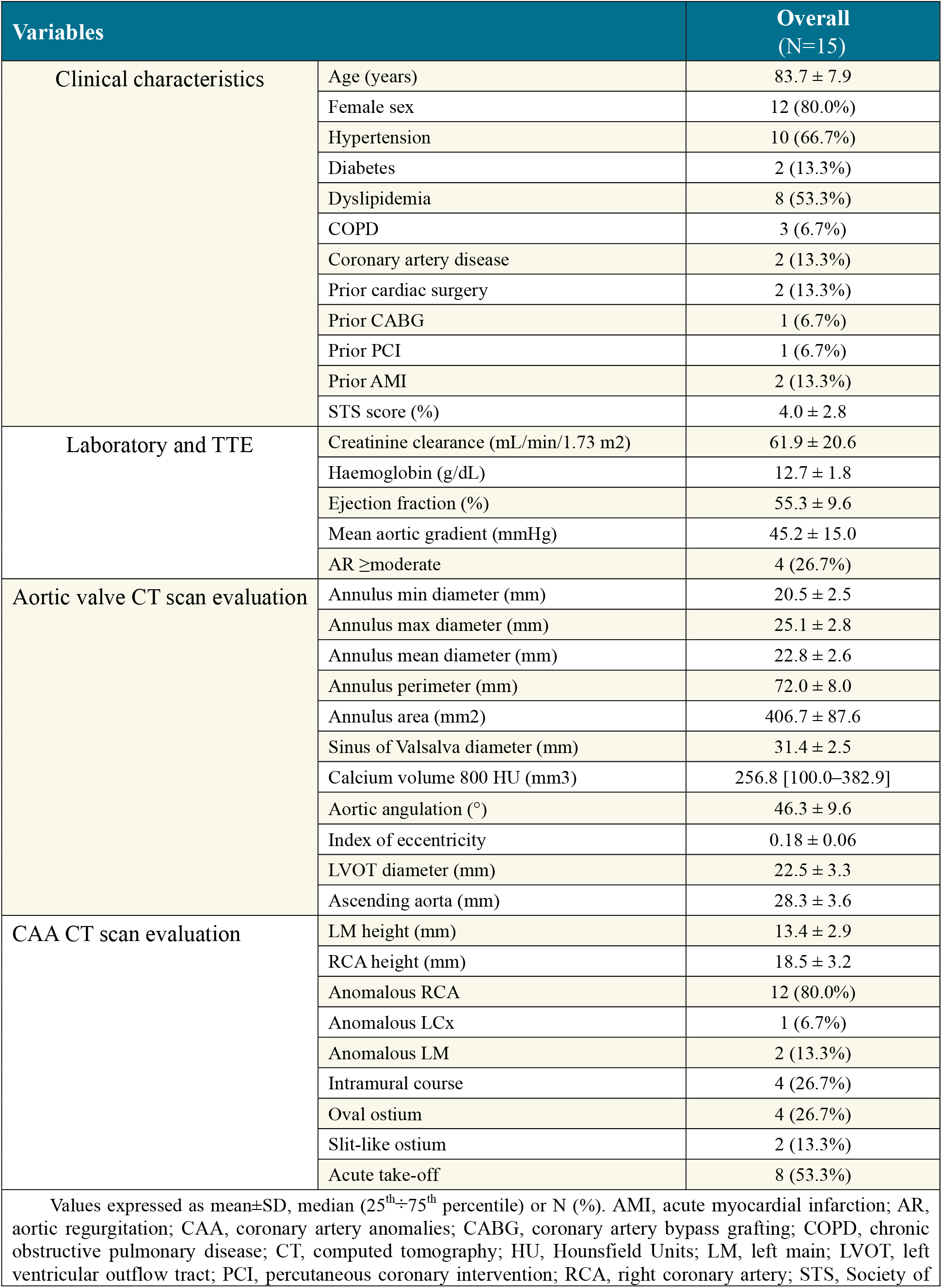
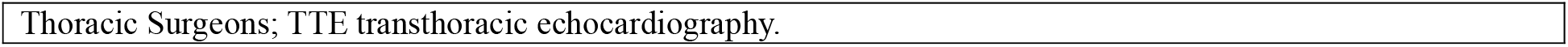
Baseline characteristics of patients with CAA who underwent TAVR.

| Variables |  | Overall<br>(N=15) |
| --- | --- | --- |
| Clinical characteristics | Age (years) | 83.7 ± 7.9 |
|  | Female sex | 12 (80.0%) |
|  | Hypertension | 10 (66.7%) |
|  | Diabetes | 2 (13.3%) |
|  | Dyslipidemia | 8 (53.3%) |
|  | COPD | 3 (6.7%) |
|  | Coronary artery disease | 2 (13.3%) |
|  | Prior cardiac surgery | 2 (13.3%) |
|  | Prior CABG | 1 (6.7%) |
|  | Prior PCI | 1 (6.7%) |
|  | Prior AMI | 2 (13.3%) |
|  | STS score (%) | 4.0 ± 2.8 |
| Laboratory and TTE | Creatinine clearance (mL/min/1.73 m<sup>2</sup>) | 61.9 ± 20.6 |
|  | Haemoglobin (g/dL) | 12.7 ± 1.8 |
|  | Ejection fraction (%) | 55.3 ± 9.6 |
|  | Mean aortic gradient (mmHg) | 45.2 ± 15.0 |
|  | AR ≥moderate | 4 (26.7%) |
| Aortic valve CT scan evaluation | Annulus min diameter (mm) | 20.5 ± 2.5 |
|  | Annulus max diameter (mm) | 25.1 ± 2.8 |
|  | Annulus mean diameter (mm) | 22.8 ± 2.6 |
|  | Annulus perimeter (mm) | 72.0 ± 8.0 |
|  | Annulus area (mm<sup>2</sup>) | 406.7 ± 87.6 |
|  | Sinus of Valsalva diameter (mm) | 31.4 ± 2.5 |
|  | Calcium volume 800 HU (mm<sup>3</sup>) | 256.8 [100.0–382.9] |
|  | Aortic angulation (°) | 46.3 ± 9.6 |
|  | Index of eccentricity | 0.18 ± 0.06 |
|  | LVOT diameter (mm) | 22.5 ± 3.3 |
|  | Ascending aorta (mm) | 28.3 ± 3.6 |
| CAA CT scan evaluation | LM height (mm) | 13.4 ± 2.9 |
|  | RCA height (mm) | 18.5 ± 3.2 |
|  | Anomalous RCA | 12 (80.0%) |
|  | Anomalous LCx | 1 (6.7%) |
|  | Anomalous LM | 2 (13.3%) |
|  | Intramural course | 4 (26.7%) |
|  | Oval ostium | 4 (26.7%) |
|  | Slit-like ostium | 2 (13.3%) |
|  | Acute take-off | 8 (53.3%) |

Values expressed as mean±SD, median (25<sup>th</sup>–75<sup>th</sup> percentile) or N (%). AMI, acute myocardial infarction; AR, aortic regurgitation; CAA, coronary artery anomalies; CABG, coronary artery bypass grafting; COPD, chronic obstructive pulmonary disease; CT, computed tomography; HU, Hounsfield Units; LM, left main; LVOT, left ventricular outflow tract; PCI, percutaneous coronary intervention; RCA, right coronary artery; STS, Society of
Thoracic Surgeons; TTE transthoracic echocardiography.

### Procedural data and outcomes

All the procedures were performed through trans-femoral approach and in 3 cases the procedure was performed with an embolic cerebral protection system. Rate of predilatation was 66.7%, meanwhile postdilation was performed only in 20% of patients. Three patients received a balloon expandable valve (BEV). Only one (ID 2) patient required femoral artery stenting due to main access vascular complication (***Table 2 and Suppl Table 2***).

**Table 2.** Procedural and follow up data of patients with CAA who underwent TAVR.

| Variables |  | Overall<br>(N=15) |
| --- | --- | --- |
| Procedure | Femoral route | 15 (100.0%) |
|  | Subclavian route | 0 (0.0%) |
|  | Embolic protection system | 3 (20.0%) |
|  | Any vascular complications | 1 (6.7%) |
|  | PTA with stenting of access site | 1 (6.7%) |
|  | PCI with stenting | 1 (6.7%) |
|  | Predilatation | 10 (66.7%) |
|  | Postdilatation | 3 (20.0%) |
|  | Emergent cardiac surgery | 0 (0.0%) |
|  | Need for second valve | 0 (0.0%) |
|  | Contrast volume (mL) | 173.8 ± 57.8 |
|  | Fluoroscopy time (min) | 21.2 ± 9.8 |
| Outcomes | Ejection fraction (%) | 56.6 ± 11.1 |
|  | Mean gradient (mmHg) | 8.2 ± 4.4 |
|  | PVL absent/trivial | 5 (33.3%) |
|  | PVL mild | 10 (66.7%) |
|  | PVL >moderate | 0 (0.0%) |
|  | Technical success | 13 (86.7%) |
|  | Device success (at 30 days) | 12 (80.0%) |
|  | PPI | 1 (6.7%) |
|  | Stroke (including not disabling) | 0 (0.0%) |
|  | 30-day mortality | 0 (0.0%) |
|  | 1-year mortality | 1 (11.1%) |
| Values expressed as mean±SD, median (25<sup>th</sup>÷75<sup>th</sup> percentile) or N (%). LCC, left coronary cusp; NCC, non-coronary cusp; PCI, percutaneous coronary intervention; PPI, permanent pacemaker implantation; PTA, percutaneous transluminal angioplasty; PVL, paravalvular leak. |  |  |

Minimal oversizing was achieved, with an overall median oversizing of 11% [5%-17%].

One patient (ID 11) experienced cardiac arrest 15 minutes after the end the procedure, due to anomalous RCA occlusion (height 14 mm). The phenotype was a CAA originating from a commissure and necessitated bailout emergency coronary artery stenting with Impella (®Abiomed) mechanical ventricular assist device. Of note, the CT scan finding was underestimated due to lack of data: a high frame valve was implanted without coronary protection and with 5 mm implantation depth, postdilatation was performed and no final selective CAA angiography was conducted.

***Table 3*** shows which valve and size has been used for each CAA phenotype.

**Table 3.** Data of CAA features and TAVR prosthesis choice.

| Patient ID | Anomalous CA | Anomalous origin | Course | Intramural tract | TAVR prosthesis | Size |
| --- | --- | --- | --- | --- | --- | --- |
| 1 | LCx | RSV | Retro-aortic | No | Portico | 25 |
| 2 | RCA | LSV | Inter-arterial | Yes | Myval | 26 |
| 3 | RCA | LSV | Inter-arterial | No | Acurate Neo | M |
| 4 | RCA | LSV | Inter-arterial | No | Evolut-R | 26 |
| 5 | RCA | LSV | Inter-arterial | No | Acurate Neo | S |
| 6 | RCA | Above STJ | Inter-arterial | No | Acurate Neo | S |
| 7 | RCA | RCC-LCC commissure | Inter-arterial | No | Evolut-R | 26 |
| 8 | LM | NCC-LCC commissure | Retro-aortic | No | Myval | 27,5 |
| 9 | RCA | Above STJ | Inter-arterial | No | Evolut-R | 26 |
| 10 | RCA | LSV | Inter-arterial | Yes | Myval | 23 |
| 11 | RCA | RCC-NCC commissure | Inter-arterial | No | Evolut-PRO | 29 |
| 12 | RCA | LSV | Inter-arterial | Yes | Navitor | 29 |
| 13 | RCA | LSV | Inter-arterial | Yes | Evolut-R | 34 |
| 14 | LM | RSV | Retro-aortic | No | Portico | 23 |
| 15 | RCA | RCC-LCC commissure | Inter-arterial | No | Portico | 25 |

CAA, coronary artery anomalies; LCC, left coronary cusp; LCx, left circumflex artery; LM, left main LSV, left sinus of Valsalva; NCC, non-coronary cusp; RCA, right coronary artery; RCC, right coronary cusp; RSV, right sinus of Valsalva; STJ, sino-tubular junction

Technical success was (86.7%), due to a cardiac arrest and a main access vascular complication, whereas device success at 30 days was lower (80%), due to the two cases of technical unsuccess and one case of aortic valve mean gradient >20 mmHg. There was no evidence of ≥ moderate PVL and of cerebral strokes. One patient deceased 4 months after the procedure, due to non-cardiac related causes (***Table 2***).

## Discussion

CAAs are rare but increasingly recognized by cardiac imaging, and a wide spectrum of alteration can be observed.^20^ The phenotypes observed in elderly patients undergoing TAVR are usually benign, even if CAAs show high-risk features.^21^

The main findings of this study can be summarized as follows:

1. the incidence of a CAA in patients undergoing TAVR is rare, below 1% of the cases;
2. myocardial infarction represents a major concern in patients with CAA undergoing TAVR as having an incidence of 6.7%: a meticulous risk assessment using CT scan is imperative.

To the best of our knowledge, this is the largest cohort of patients with a CAA undergoing TAVR, and a number of technical considerations can be done based on the cases performed; as such, we hereby propose a classification, based on phenotypes of CAA, and two algorithms aimed at preventing possible CAA related complications.

The first group is composed by patients deemed “at risk of ab extrinseco CAA compression”, due to valve implantation (***Central Illustration A***). In this subset of patients, occlusion of coronary arteries with a retroaortic course is the most common complication described in literature, due to the strict anatomical relationship between the CAA and the aortic root, where the radial forces generated by the bioprosthesis are exerted ^22–25^ An interarterial course is more common and theoretically associated with a lower risk of compression due to the shorter vessel segment at risk (to be evaluated case by case)^26–28^. The intramural course can be combined with the previous ones and it is the least described in literature,^29^ probably due to the low impact on coronary artery perfusion in patients who did not show sudden cardiac death in a lifelong.

There are a few strategies described in literature to avoid CAA compression:

1. to assess the effective risk with an aortogram taken during balloon predilatation.^24,26,27^ As a matter of fact, this procedure is not performed as a routine practice, but it is reserved only for patients exhibiting high-risk features (e.g. retro-aortic course). Additionally, predilatation with selective CAA angiography is feasible;^23^ however it is technically more demanding and entails the risk of proximal vessel injury.
2. the use of self-expandable valves (SEV) which have the capability to be recaptured, thus allowing for valve removal in cases of relevant overt compression.^24,29,30^ In our study, recapturable devices were used in 9 out of 15 patients: low radial force SEV,^31^ such as the Portico^32^ and Acurate Neo, have the advantage of exerting lesser impact on the aortic root with minimal deformation. In our series, low radial force SEV were utilized in 6/15 patients.
3. the use of BEV that, unlike SEV, do not keep generating forces on the aortic root after deployment.^27^ BEV was used in 3 out of 15 patients.
4. Bioprosthesis undersizing allows lower stress and aortic anulus deformation. Oversizing in high-risk features is always not recommended,^22^
5. coronary protection with the chimney technique, especially when there is evidence of CAA obstruction during balloon valvuloplasty, in order to quickly deal with the abrupt coronary occlusion.^33^ In our series it was performed in 2/15 patients.

The second group is composed by patients deemed “at risk of CAA ostium occlusion” (***Central Illustration B***). Historically, CAA originating from the commissure between two cusps have not been considered at risk for coronary occlusion: this is true when the aortic root is large enough to accommodate the bioprosthesis and still there is enough space between the latter and the coronary ostium. This measure is typically evaluated while planning a VIV procedure, i.e. the “VTC: valve-to-coronary”. Of note, there is no standard reference to consider in case of a CAA ^33,34^

In our sample, 1 patient over 4 experienced an acute CAA occlusion. Consequently, we propose the following strategies:

1. mild “intentional” misalignment, to avoid placing a strut in front of the CAA ostia.^33^ In our series the only valve that could be deliberately misaligned were the Acurate Neo and Evolut. However, being a retrospective study, it was not systematically used.
2. Low frame valve deployment (BE might be appropriate), to lessen the interaction of the upper crown with leaflets and CAA ostium (1/4 patients).
3. A slightly deeper implantation, to reduce leaflets eversion although it may increase the risk of conduction disturbances.^35^
4. Coronary protection thorough the chimney technique, as described previously. We suggest performing this technique in anatomies with highest risk of occlusion.

Additional general strategies for these two groups are: i) avoidance of postdilatation, if feasible (20% patients in our sample), in order to avoid further stress on the aortic root and eventual compression and ii) final selective CAA angiography^31,34^ to verify patency of the CAA (3/15 in our sample).

The final group consists of patients at “high risk of impaired CAA re-access”. This group encompasses (a) CAA located above the sino-tubular junction (STJ), as well as (b) CAA originating from the commissure and (c) from the opposite sinus of Valsalva (***Central Illustration C***) where the distance is enough to make an abrupt occlusion of the coronary artery unlikely but still the cannulation might be cumbersome. For the latter two types, the suggested strategies for preventing CAA ostium occlusion remain the same. However, there are two main strategies that we suggest to preserve coronary re-access in a CAA above the STJ:

1. use of a valve with a lower height than the origin of the CAA (ID 9: in this case it was not applied due to the patient’s age of 90 years old and no coronary artery disease),
2. and use of an open frame valve, as the Acurate Neo (ID 6). Both strategies are intended to avoid the interference of the upper crown with catheters.

To systematically evaluate and manage the possible consequences of the presence of a CAA during TAVR, we hereby present two algorithms.

The first algorithm is based on the mechanism of CAA flow impairment, and it is described in ***Figure 1***. Firstly, if the patient belongs to group 1 (risk of ab extrinseco compression), balloon predilatation with concomitant angiography should be performed. The subsequent strategy depends on the presence of compression. If the patient is in group 2 (risk of coronary ostium occlusion), the strategies will be different according to the type of CAA (commissural origin vs. low origin from opposite sinus of Valsalva). General strategies for both groups are avoidance of postdilatation and final selective CAA angiography.

**Figure 1.**
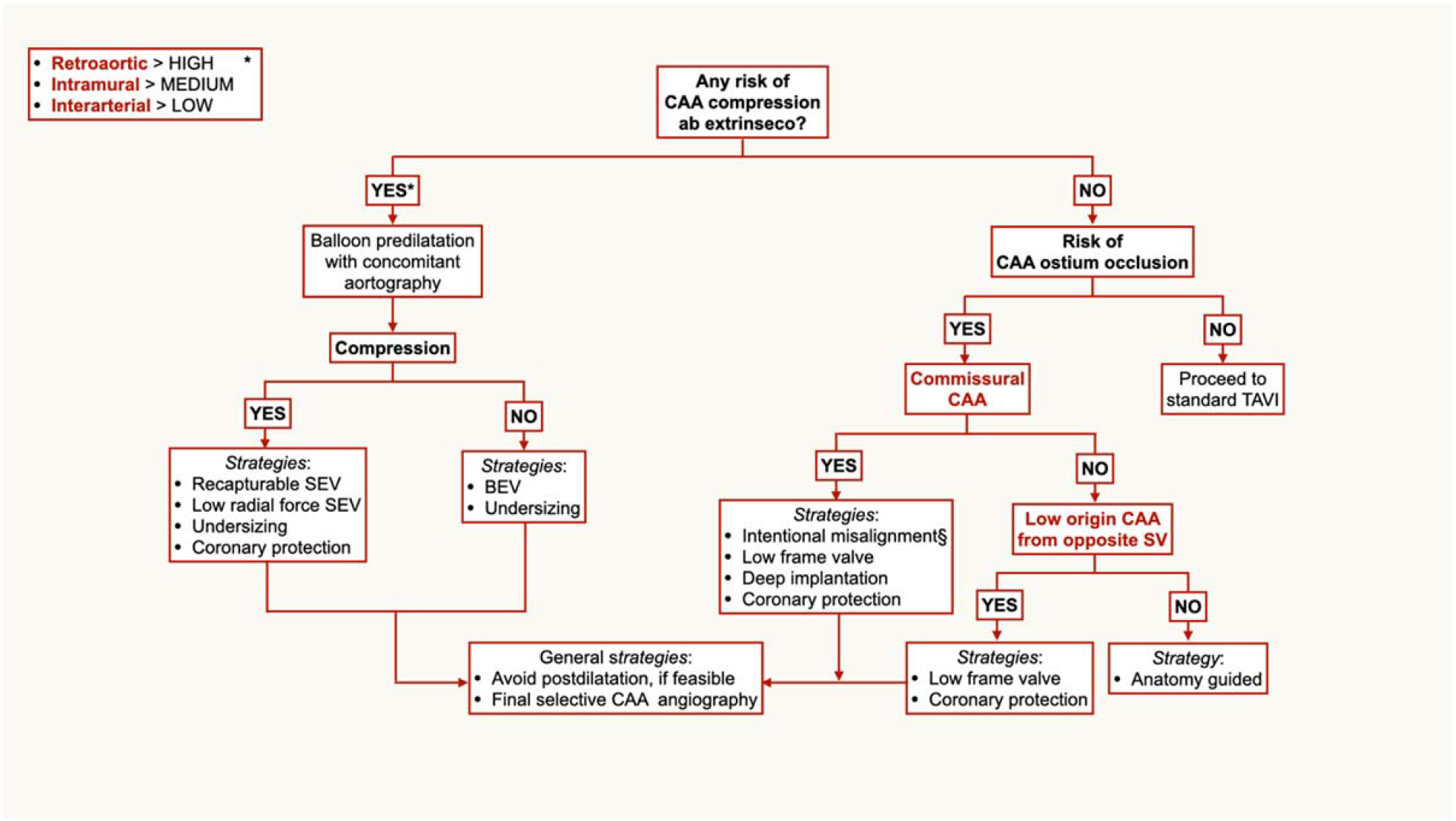
Algorithm for valve choice in patient with CAA undergoing TAVR, at risk of coronary occlusion. *Retroaortic, intramural and interarterial CAA have different risk of coronary compression and subsequent occlusion, but the anatomy must be assessed case-by-case. However, the algorithm is not affected, due to data paucity and unpredictable behavior in real world. §Placing of a neocommissure in front of the CAA ostium, through a custom projection, detected by CT scan. BEV, balloon-expandable valve; CAA, coronary artery anomaly; SEV, self-expandable valve; SV, sinus of Valsalva.

The second algorithm is based on the risk of impaired coronary re-access (group 3), as shown in ***Figure 2***. Strategies are different based on the phenotype: origin above the STJ, commissural origin and low origin from opposite sinus of Valsalva.

**Figure 2.**
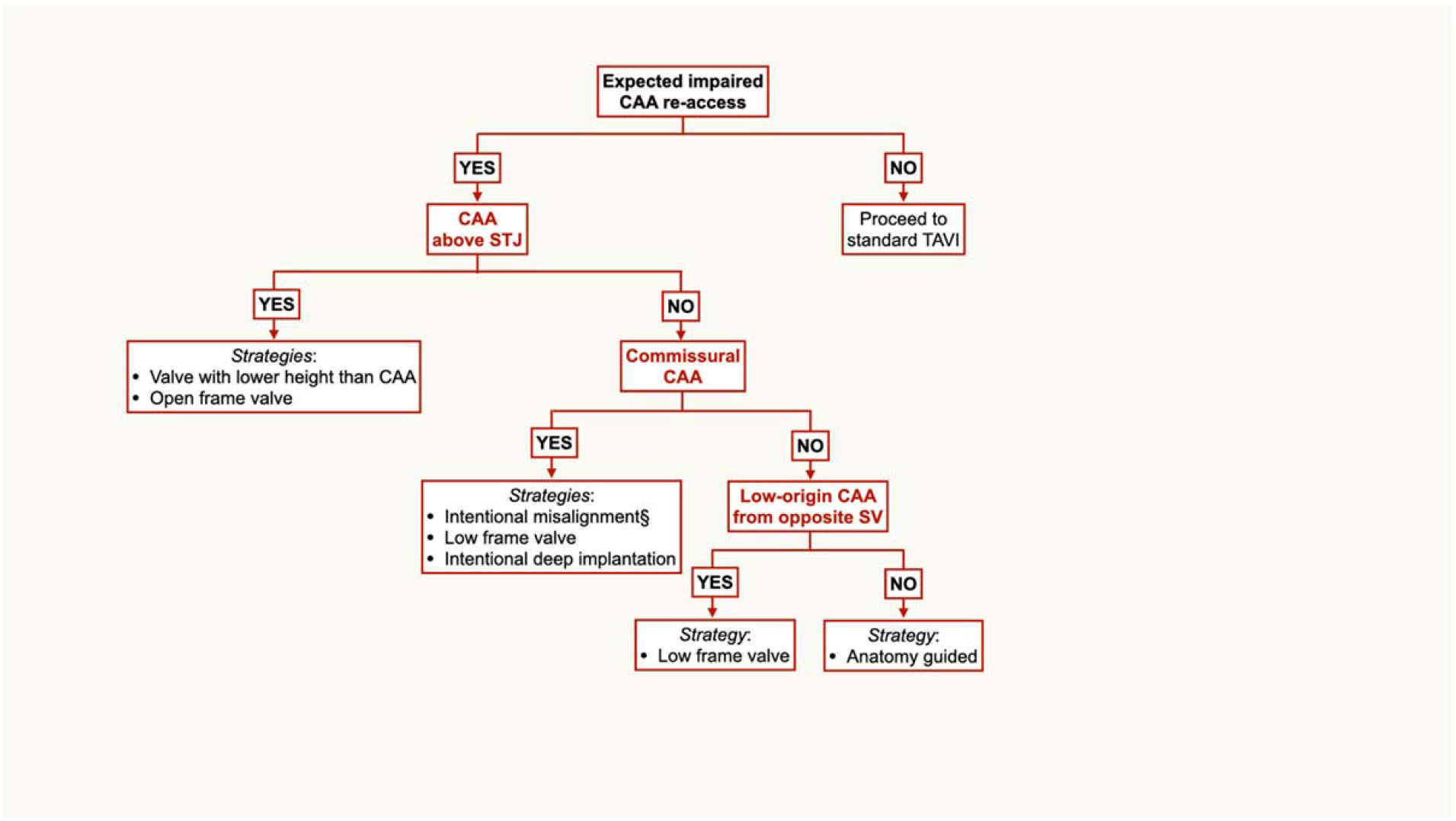
Algorithm for valve choice in patient with CAA who undergo TAVR, at risk of impaired coronary re-access. §Placing of a neocommissure in front of the CAA ostium, through a custom projection, detected by CT scan. CAA, coronary artery anomaly; STJ, sino-tubular junction; SV, sinus of Valsalva.

## Study limitations

Data analysis has been conducted retrospectively and the small number of patients did not allow more extensive data investigation.

Due to CAA rarity and their heterogeneity, the results of the analysis still must be considered exploratory; further studies applying the proposed algorithms are needed for a thorough validation.

## Conclusions

The presence of a CAA may significantly increase the risk of threatening complications during TAVR. The possible interaction between the transcatheter bioprosthesis and the different CAA types should be comprehensively evaluated upfront and the procedure planned accordingly.

## Data Availability

The authors confirm that the data supporting the findings of this study are available within the article [and/or] its supplementary materials.

## Abbreviations list

BEV: balloon-expandable valve
CAA: coronary artery anomalies
CT: Computed Tomography
LAD: left anterior descending
LCx: left circumflex artery
LSV: left sinus of Valsalva
PVL: paravalvular leak
RCA: right coronary artery
RSV: right sinus of Valsalva
SAVR: surgical aortic valve replacement
STJ: sino-tubular junction
TAVR: transcatheter aortic valve replacement
TTE: transthoracic echocardiography
VARC-3: Valve Academic Research Consortium-3
SEV: self-expanding valve.

## Acknowledgements

None

## Figure Titles and Legends

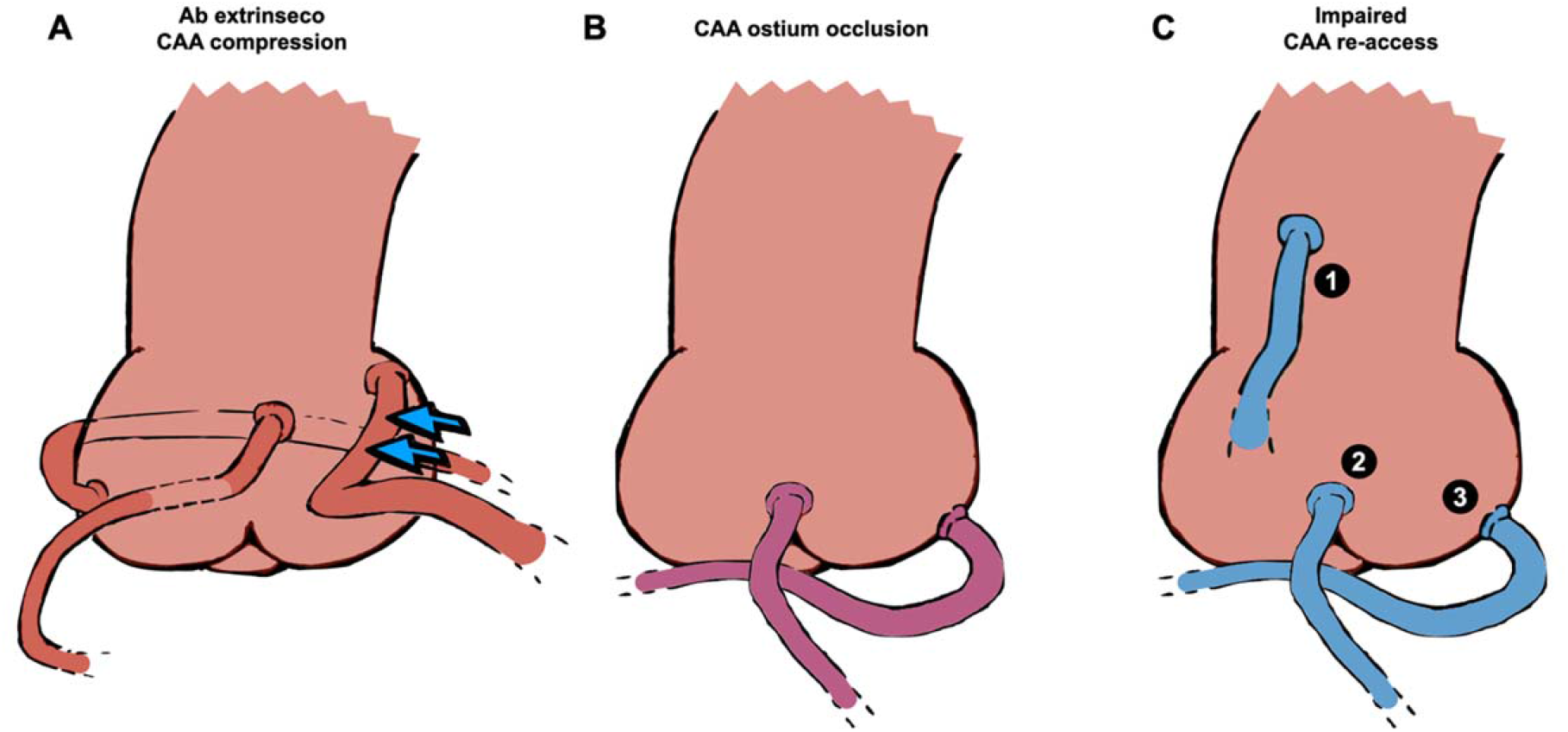
Central Illustration. Most common phenotypes of CAA in patients undergoing TAVR, classified by mechanism of impairment. (A) Compression ab extrinseco (red). From left to right: retroaortic course, intramural course and interarterial course (blue arrows to represent pulmonary artery compression). (B) Ostium occlusion (purple). From left to right: commissural origin and low origin from opposite SV. (C) Challenging re-access (blue). 1) above STJ origin; 2) commissural origin and 3) low origin from opposite SV.

